# The Effects of Expectations and Worries on the Experience of COVID-19 Symptoms

**DOI:** 10.1101/2023.01.26.23284911

**Authors:** Titilola Akintola, Joyce Chung, Lauren Atlas

## Abstract

**Introduction:** The COVID-19 pandemic has been shown to have profound effects on both mental and physical health. Distress and widespread uncertainty about global events and personal risk are associated with increased worry and negative expectations that impact physical health. Thus, the current pandemic poses a possibility for the experience of nocebo effects.

**Objective:** To evaluate the likelihood of nocebo-induced COVID-19 symptoms in a US sample.

**Methods:** An online study on the mental health impact of COVID-19 asked participants to complete a set of biweekly surveys over a 6-month period between April 2020 and May 2021. We focus on responses from 3,027 individuals who reported never testing positive for COVID-19. We assessed the association between two types of worry and self-reported symptoms of COVID-19. We used multi-level models to examine variations across and within participants over time. We further investigated the effects of pre-existing health conditions and mental health status.

**Results:** There was a positive association between symptoms and both general (b= 2.56, p<0.01) and personal worry (b=2.77, p<0.01). However, worry reported at one timepoint was not specifically associated with symptoms reported two weeks later (p = 0.63, p=0.56). We also found that a greater number of prior clinical comorbidities and greater mental health burden were significant predictors of symptom reporting.

**Conclusions:** These results suggest that increased worries during the COVID-19 pandemic were associated with greater symptoms. Further studies investigating worry and symptoms in populations with confirmed negative COVID-19 tests or isolated populations will be needed to isolate the occurrence of true nocebo effects during the pandemic.

## INTRODUCTION

The 2019 Coronavirus Disease (COVID-19) is arguably one of the greatest public health challenges of this millennium, reaching every continent and causing far-reaching implications to every aspect of modern life. The combination of significant changes to society and lifestyle, quarantine measures, and uncertainty about the future has positioned the COVID-19 as a major stressor with extensive psychosocial impact, similar to previous pandemics ((Whitehead 2020, Esterwood and Saeed, 2020). One specific effect of the COVID-19 pandemic has been wide-spread increases in distress, hopelessness and worries about personal and global outcomes (Dubey, Biswas et al. 2020, Tull, Edmonds et al. 2020). Specifically, worry about health has been understandably reported to increase (Taylor, Asmundson 2004; Daniali 2021) and spread during past pandemics such as the Ebola and H1N1 Influenza viruses (Bish and Michie 2010; Blakey 2015; Xie 2011).

Particularly, with this current pandemic, the greater access to the internet coupled with constant changing or conflicting information from various sources, all contribute to even greater stress and more downstream effects (Amanzio, 2020). Studies have shown that affective factors such as worry and hope can be linked to the generation of self-directed expectations that can influence personal well-being and health outcomes (Hirsh 2015, Di Blasi 2001, El-Haddad 2020, Wiles, Cott et al. 2008).

Previous research investigating the association between expectations and health outcomes indicate that changes in expectations can be linked to changes in clinical symptoms, pain, disability and other health outcomes (Kirsch 1985, Finniss, Kaptchuk et al. 2010). For example, studies show that positive expectations are associated with better health outcomes (Myers, Phillips et al. 2008, Iles, Davidson et al. 2009, Eklund, De Carvalho et al. 2019) while more negative expectations and contextual factors may result in worse health outcomes – in line with the nocebo phenomenon (Hahn 1997, Colloca 2011; Hauser 2012; Bingel 2014).

Nocebo effects can be described as adverse health symptoms or events that occur due to negative expectations and contextual factors. Thus, during the current pandemic it is also likely that worries and health anxiety will be increased, possibly leading to adverse health effects or nocebo effects. The likelihood of this is also amplified by continuously changing information in the media, the abundance of conspiracy theories, and uncertainty (Amazio 2020).

In fact, a few studies investigating COVID-19 vaccine hesitancy and the experience of adverse effects report that fear and negative expectations may have induced adverse effects in vaccine trials (Polack 2020), consistent with nocebo effects. A cross-sectional study of over 25,000 participants in France showed that *beliefs* about having had a COVID-19 infection were associated with increased self-reports of multiple, persistent COVID-19 symptoms (long-COVID) while actual positive laboratory serology tests were only associated with anosmia (Matta 2021). Given these findings, it is important to assess how expectations and worry may influence the experience of COVID-19 symptoms and other clinical and psychological health outcomes during the ongoing COVID-19 pandemic. This is especially important in more vulnerable populations such as those with mental health conditions or long-term chronic conditions who may be susceptible to experience compounding psychophysical effects due to the potential interaction of these conditions (Amerio 2020), such as some in our study population. Although the effects of the ongoing pandemic on mental and physical health is still being unraveled, the current context supports a framework conducive for possible nocebo effects (Benedetti 2007, Amanzio 2020).

Motivated by these well-documented findings on placebo and nocebo effects, we explored the association between worry and COVID-19 symptoms during the pandemic. We tested the pre-registered hypothesis that worry during the coronavirus pandemic would be associated with the experience of COVID-19 symptoms in a longitudinal study of over 3,500 individuals during the first year of the pandemic (https://aspredicted.org/blind.php?x=42i5xr). We hypothesized that the experience of COVID-19 symptoms would be influenced by both external worry about others or global outcomes (general worry) and self-directed worries about personally becoming infected (personal worry), and that the association between general worry and symptoms would be mediated by worries about personal health. We tested these hypotheses across individuals and asked whether individuals who reported greater worries also reported greater symptoms. To address the likely bidirectionality of these associations at any given timepoint (i.e., a participant’s worry could influence COVID-19 symptoms experienced and the experience of symptoms could also modulate the worry), we complemented our between-participants analysis with within-subjects time-lagged analysis to test directional hypotheses. More specifically, we tested the hypothesis that general and personal worry at one time point would be positively associated with physical symptoms at the next time point.

## METHODS

### Recruitment & Sample Population

The over-arching project titled “Mental Health Impact of COVID-19 Pandemic on NIMH Research Participants and Volunteers” was part of a collaborative effort involving multiple National Institute of Mental Health Intramural Research Program (NIMH IRP) investigators and affiliates. The project was launched in April 2020 in response to the emergent COVID-19 pandemic and aimed to examine the potential effects of stressors related to the pandemic on various mental, physical health and behavioral factors (Chung 2021). Other general details of the study design and objective are available online at https://clinicaltrials.gov/ct2/show/NCT04339790.

This study was conducted completely online, and participants were recruited both from a pool of study participants previously enrolled to several NIMH IRP protocols as well as from the general populace. Recruitment was carried out via invitations / emails, flyers, social media ads, postings on listservs and clinicaltrial.gov as well as via word of mouth. Official enrollment took place from April 4 through November 1, 2020. Eligibility for this study was limited to English-speaking adults, aged 18 or older. A total of 3,655 participants enrolled in the study with representation from all 50 U.S. states and some (∼1%) international participants. While all participants were asked to complete surveys every two weeks, some missed intervals during the 6-month follow-up period. To ensure our average calculations captured multiple timepoints and not one isolated occurrence, we excluded participants who only provided survey responses for a single timepoint from this analysis. In addition, we excluded participants who reported testing positive for COVID-19 at any point during the study.

### Study Procedures

#### Baseline Procedures

Following online consent, each participant filled out enrollment survey measures including demographics, the DSM-5 Self-Rated Level 1 Cross-Cutting Symptom Measure-Adult (Bastiaens, 2018) as well as clinical and mental health history questionnaires. Participants were also asked to complete the Psychosocial Impact of COVID-19 survey (COVID-19 survey), developed for the study, which includes several COVID-19-related outcomes and the 3-item UCLA Loneliness Scale (Russel, 1996) (full survey available at https://tools.niehs.nih.gov/dr2/index.cfm/resource/22587). For the present analysis, we focus on questions from the COVID-19 survey about worry and expectations related to COVID-19, participants’ COVID-19 testing and results, demographics and self-reports of any COVID-19 associated symptoms as discussed in more detail below. For a complete list of measures collected in the broader survey please see Chung et al., 2021.

#### Biweekly Procedure

Following enrollment, participants were asked to complete a set of online surveys which included the COVID-19 survey every two weeks for a period of 6 months (24 weeks, or 12 intervals)from their enrollment timepoint.

#### End of Study Procedures

At the final timepoint (12^th^ interval), participants were asked to complete several additional measures, including a Chronic Pain Graded Scale (CPGS) (Von Korff 1992) to determine the presence and intensity of Chronic Pain.

#### Study Measures

Details about each outcome measure used for these analyses and how they were operationalized are as follows:

##### Worry Measures

The COVID-19 survey included multiple questions regarding different sources of worry. For the purpose of this paper, we focused on survery items (7 -12 & 37) of our survey, which center on worries about the (COVID-19) pandemic in general, worry about family members becoming infected, worries about others becoming infected, worry about access to food, worries about access to other resources, worries about personally becoming infected with the coronavirus and worries about personal physical health being affected. For each measure, participants were asked to report how worried they were on a scale of 1 (not at all worried) to 10 (extremely worried). Based on our hypotheses about general or external worry and self-directed worry, we operationalized worry into two measures: “general worry” and “personal worry”. We anticipated that the responses to some of the worry measures might be correlated, so we performed Pearson’s correlations to determine which to classify as general worry or personal worry. There was a strong (all r’s > 0.7) and significant (all p’s < 0.01) correlation between participants’ worries about the pandemic, worries about family health and worries about others; thus at each timepoint “general worry” was computed as the mean of the following three items for each participant: : “How worried are you about coronavirus (COVID-19)?” (survey item 7); “How worried are you that a family member would be infected with coronavirus (COVID-19)?” (survey item 9); “How worried are you that others around you will be infected with coronavirus (COVID-19)?” (survey item 10). Similarly, participant’s worries about becoming infected and worries about their physical health being affected were highly correlated with each other (r=0.87, p<0.01), thus a mean of the following responses at each time point was computed as “personal worry”: “How worried are you that you will be infected with coronavirus (COVID-19)?” (survey item 8); *“How worried were you that your physical health could be affected by the coronavirus (COVID-19) pandemic?”* (survey item 37) (see Table 1). Worry about access to food and access to other resources were strongly correlated with each other (r = 0.8, p <0.01) but were weakly correlated (r < 0.20) with all other worry measures and so were not included.

**Table 1.a.** Pearson Correlations of General Worry Measures.

| Table 1.a Pearson Correlations of General Worry Measures |  |  |  |  |  |
| --- | --- | --- | --- | --- | --- |
|  | 1 | 2 | 3 | 4 | 5 |
| 1. Worry about the pandemic | - | 0.798*** | 0.845*** | 0.207*** | 0.169*** |
| 2. Worry about family members being infected | 0.798*** | - | 0.860*** | 0.166*** | 0.137*** |
| 3. Worry about others being infected | 0.845*** | 0.860*** | - | 0.192*** | 0.156*** |
| 4. Worry about access to food | 0.207*** | 0.166*** | 0.192*** | - | 0.819*** |
| 5. Worry about access to transportation or housing | 0.169*** | 0.137*** | 0.156*** | 0.819*** | - |
| r values displayed |  |  |  |  |  |
| ***Correlation is significant (p < 0.01) (2-tailed) |  |  |  |  |  |

**Table 1.b.** Pearson Correlation of Personal Worry Measures.

| Table 1.b Pearson Correlation of Personal Worry Measures |  |  |
| --- | --- | --- |
|  | 1 | 2 |
| 1. Worry about being infected | - | 0.873*** |
| 2. Worry about physical health being affected | 0.873*** | - |
| r values displayed |  |  |
| ***Correlation is significant (p < 0.01) (2-tailed) |  |  |

##### Symptoms

At each interval, participants reported whether they had experienced any of the common symptoms of COVID-19. The initial questionnaire asked participants to report if they had experienced any of the following: congestion, runny nose, sore throat, cough, fever, headache, fatigue, diarrhea and shortness of breath. As more knowledge of the COVID-19 sequelae became available, we included the following additional symptoms: chills, shaking with chills, muscle pain, new loss of taste, new loss of smell or other. Taking these changes into account, at each timepoint we computed a proportion of symptoms reported by participants out of the total number of symptoms (9 or 15) available. We then calculated an average proportion of symptoms reported throughout the survey period for analyses across individuals, whereas analyses within individuals measured the proportion of symptoms reported at a given timepoint.

##### Comorbidities

At baseline, we collected participants’ self reported prior clinical history and from this computed the total sum of prior comorbidities for each participant. Our clinical history form asked participants to indicate if they had a personal history of cancer, heart disease, high blood pressure, diabetes, stroke, lung, liver, kidney or thyroid disease, a stomach condition, an immune disorder or other disorder. Each positive response to any these conditions was given a score of 1 and the total number of comorbidities per participant was included as a predictor of symptoms in subsequent analyses.

##### Mental Health Burden Assessment

Because a proportion of our study participants were pooled from past NIMH study participants and were likely to include people with mental health conditions, we included this in our model. To investigate mental health status as a predictor of COVID-19 symptom reporting, we utilized a machine-learning derived Patient Probability Score (PPS) that estimates the likelihood of having a mental health diagnosis. PPS scores were derived using study enrollment questionnaires and trained on participants who had previously been in NIMH studies that administered a mental health diagnostic assessment, the Structured Clinical Interview for DSM; (for additional details, see Chung et al., 2021). PPS provide a continuous score (0-1) for each participant’s likelihood of having a mental health diagnosis.

#### Statistical Analysis

This study included several analyses to test the relationship between worry and reported symptoms in our sample population, as discussed below. All analyses were performed in RStudio version 1.4.1106 (RStudio, Inc., Boston, MA).

##### Association between worry and symptoms across individuals

We used linear regression (*lm*) package, version 3.6.3 in RStudio version 1.4.1106 (Rstudio, Inc., Boston, MA) to test whether individuals who reported experiencing greater worry on average also reported a greater average proportion of symptoms relative to individuals who experienced less worry. Models treated average proportion of symptoms as the dependent variable and worry (general or personal) as the independent variable. Since some of our covariates were correlated, we included them separately in different models to allow for a matrix condition number (kappa) of < 3, we ran separate linear regression models: i.) controlling for age and sex and ii.) testing for interactions with number of physical comorbidities and mental health burden. All predictors included in the models were mean-centered.

We then tested whether average personal worry mediated the relationship between average general worry and average proportion of symptoms reported, using the *lavaan* R-package (Rosseel 2012). We performed the Sobel test (Sobel, 1982), using the Delta method assuming normal distribution of the sampling distribution. We used 95% percentile confidence intervals (Cis) generated by a bootstrapping procedure with a resampling rate of 1,000 to evaluate the reliability of the mediation effect.

##### Time-lagged association between worry and symptoms within individuals

To explore the presence of a directional effect of worry on symptoms, we performed within-subjects analyses using the linear mixed-effects model *nlme* package, version 3.1-144 in RStudio version 1.4.1106 (RStudio, Inc., Boston, MA). We tested the relationship between participants’ worry at each timepoint (T), on the proportion of symptoms reported at the next timepoint (T+1), conducting separate analyses for general and personal worry. For these analyses, we only included participants who submitted entries for all 13 biweekly timepoints during the survey period (n =543). To address the hierarchical nature of these analyses and test whether associations varied as a function of individuals’ overall level of worry, we modelled participants’ mean worry (general or personal) over the 13 timepoints as between subject effects (2^nd^ level variable). For both analyses (symptoms (T+1) ∼ general worry (T) and symptoms (T+1) ∼ personal worry (T)), we treated intercept and slope as random (Lindstrom & Bates, 1990, Barr 2013) and included the cluster mean centered value and the grand mean centered cluster mean value as fixed effects, testing for main effects and interactions. We used the *nlm*e function method “Maximum Likelihood” (ML) and an autocorrelation (AR1) model to account for the autocorrelation of regression residuals (Lindstrom & Bates, 1990).

## RESULTS

### Participants

A total of 3,655 participants enrolled in the study. 105 participants (2.9%) reported testing positive for the coronavirus at some point during the survey period and thus were excluded from this analysis. Furthermore, 459 participants (12.6%) completed only one timepoint (enrollment) and thus were removed from this analysis. We further excluded 64 participants for whom we didn’t have Mental Health status data on. Thus, our between subject analyses included the 3,027 participants who responded to the mental health questionnaires and the COVID-19 survey at least 2 times. 2,487 were women (∼82%) and 518 were men (∼17%), while <1% of participants did not report biological sex. The sample was also predominantly Caucasian with ∼91% self-identifying as Caucasian, while ∼3% self-identified as African-American or Black and ∼3% as Asian. The mean age was 47.17 (SD=14.8).

### Greater General Worry about the Pandemic is Associated with Increased reporting of Covid-19 Symptoms

Consistent with our first hypothesis, linear regression models revealed a main effect of average general worry on the proportion of COVID-19 symptoms reported, while controlling for sex and age (ß =2.56, p<0.01), such that those who reported greater average worry reported more symptoms on average. There was also a main effect of age (ß =-0.90, p <0.01) such that younger participants tended to report more COVID-19 symptoms, and a significant effect of sex such that females reported more symptoms (ß=0.72, p<0.01). There were no interaction effects. Taking into account the number of comorbidities and mental health status (PPS), there was still a significant relationship between average general worry and number of symptoms reported (ß =1.94, p <0.01). In addition, we observed main effects of number of physical comorbidities (ß = 1.38, p<0.01) and mental health status (ß =3.02 and p< 0.01), such that participants with greater comorbidities or mental health burden also reported greater COVID-19 symptoms. There was also a small but significant interaction effect between general worry, number of comorbidities and mental health status (ß =-0.49, p=0.02) (shown in Fig.1. a, Table 3. a). Correlation results for all variables and control variable included in the models are reported in Table 2.

**Fig 1 A:**
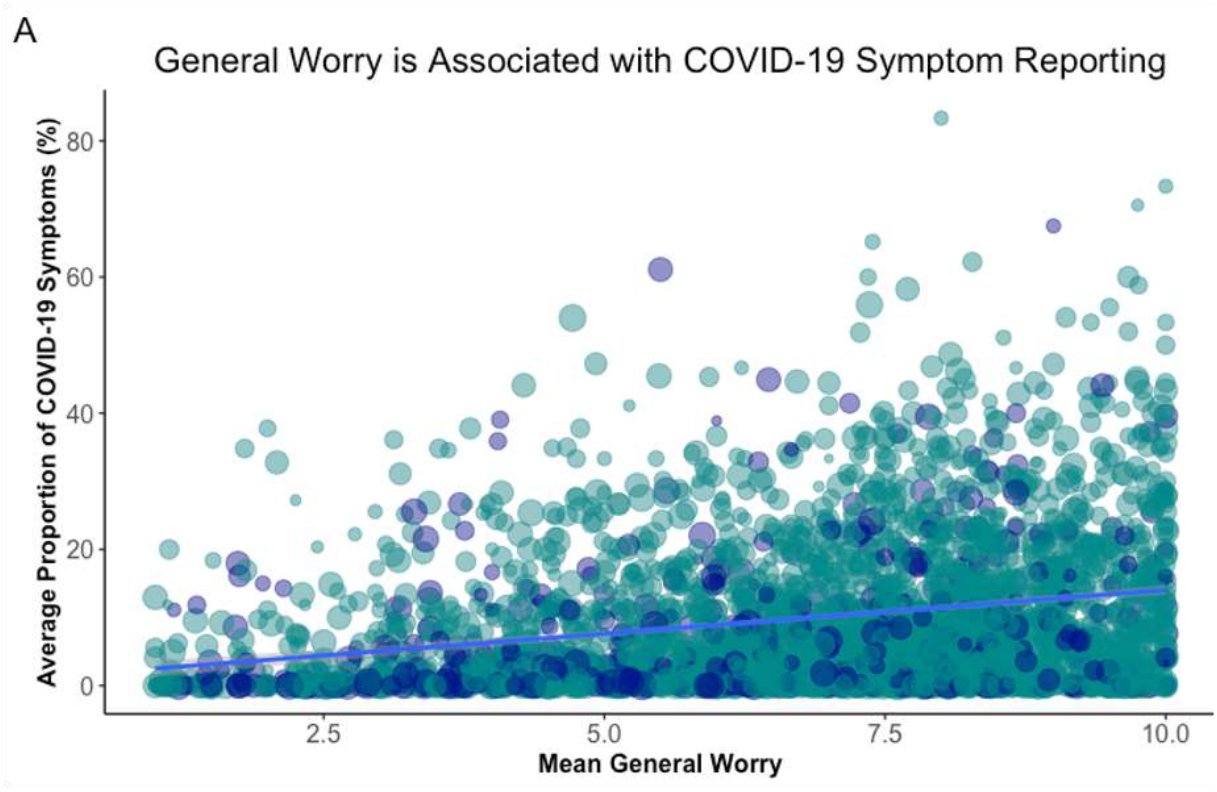
Scatter plot shows the association between Participants Mean General Worry and the average Proportion of Symptoms reported over the 6-month survey period controlling for age & sex (B = 2.56, p < 0.001, Table 3i) and including participants number of comorbidities and Mental Health Burden score as predictors (B = 1.94, p < 0.001, Table 3ii). All other interactions tested; and significant ones reported in table.

### Greater Worries about Personal Health Are Associated with Increased reporting of Covid-19 Symptoms

Consistent with hypotheses, we observed similar trends when we analyzed the association between symptoms and average worry for personal health and infection. Controlling for age and sex, average personal worry was associated with increased symptom reporting (ß = 2.77, p<0.01). Again, younger participants (ß = -1.11, p<0.01) and women (ß = 0.71, p<0.01) reported more COVID-19 symptoms There were also significant interactions between age and personal worry (ß=-0.43, p =0.04) on symptom reporting. Due to the negligible sizes of these correlations, we refrain from inferring any clinically relevant differences in the effect of personal worry on symptom reporting between younger and older participants. Taking into account the number of comorbidities and mental health burden of participants, we found still a main effect of average personal worry (ß =1.95, p<0.01), and found that symptom reporting was also positively associated with a greater number of physical comorbidities (ß =1.17, p<0.01) and mental health burden (ß =3.07, p <0.01) (shown in Fig.1. b, Table 4).

**Fig 1 B:**
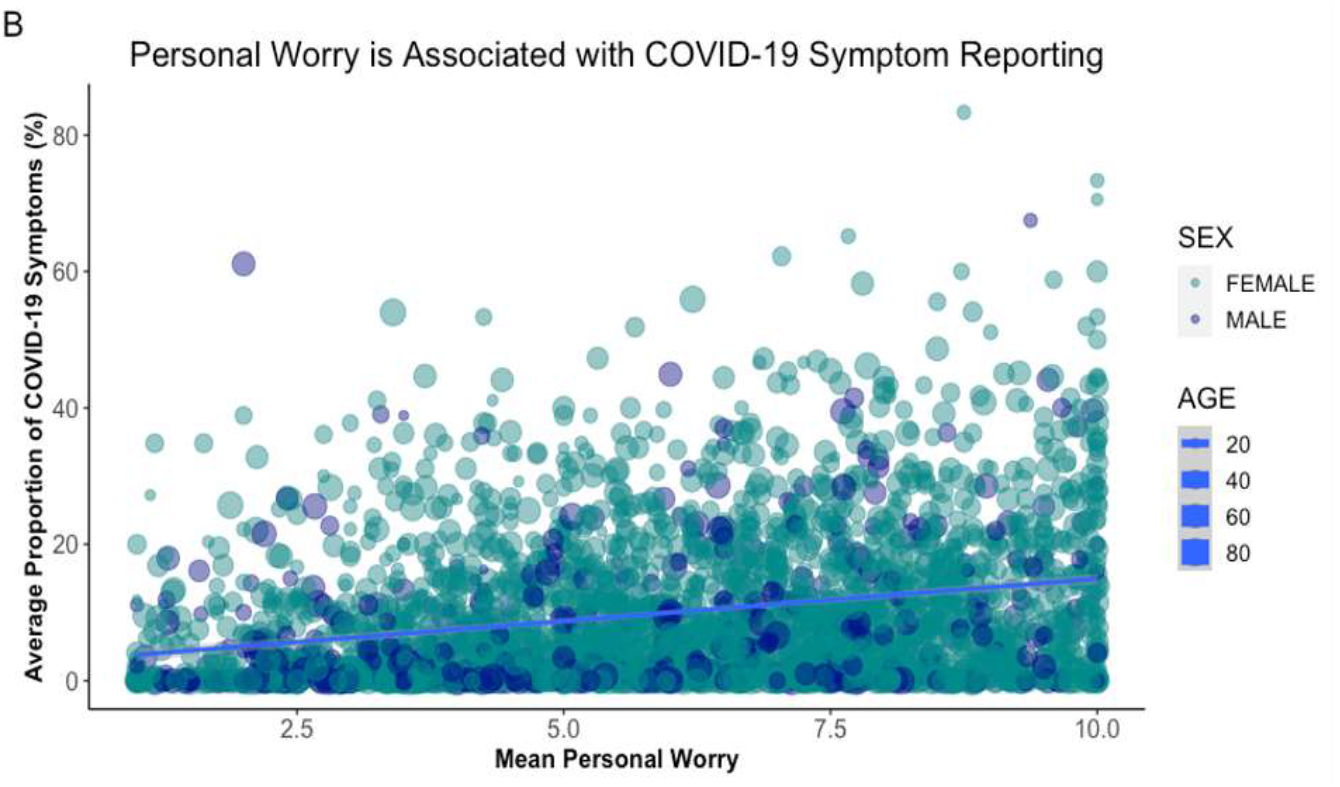
Scatter plot shows the association between Participants Mean General Worry and the average Proportion of Symptoms reported over the 6-month survey period controlling for age & sex (B = 2.77, p < 0.001, Table 4i) and including participants number of comorbidities and Mental Health Burden score as predictors (B = 1.95, p < 0.001, Table 4ii)). All other interactions tested; and significant ones reported in table.

### Personal Worry Mediates the Relationship between General Worry and symptoms

We used mediation analysis to test the hypothesis that the associations we observed between general worry and symptoms were mediated by personal worry (Figure 2). Path “a” indicated that general worry was positively associated with personal worry (ß = 0.940, SE = 0.01, 95% CI = [0.923, 0.958], p < 0.01). Path “b” indicated that personal worry was positively associated with the proportion of COVID-19 symptoms reported, controlling for the effect of general worry (ß = 0.876, SE = 0.18, 95% CI = [0.512, 1.218], p < 0.01). The indirect effect of general worry on average symptoms through the effect of personal worry was also significant (path “ab” = 0.824, SE = 0.171, 95% CI = [0.483, 1.149] p < 0.01), indicating that personal worry is a significant mediator of the association between general worry & COVID-19 symptoms. Finally, the direct effect of general worry on symptoms remained significant (path “c”: ß = 0.458, SE = 0.180, 95% CI = [0.121, 0.798], p = 0.01) suggesting a partial mediation (shown in Fig. 2).

**Fig 2.**
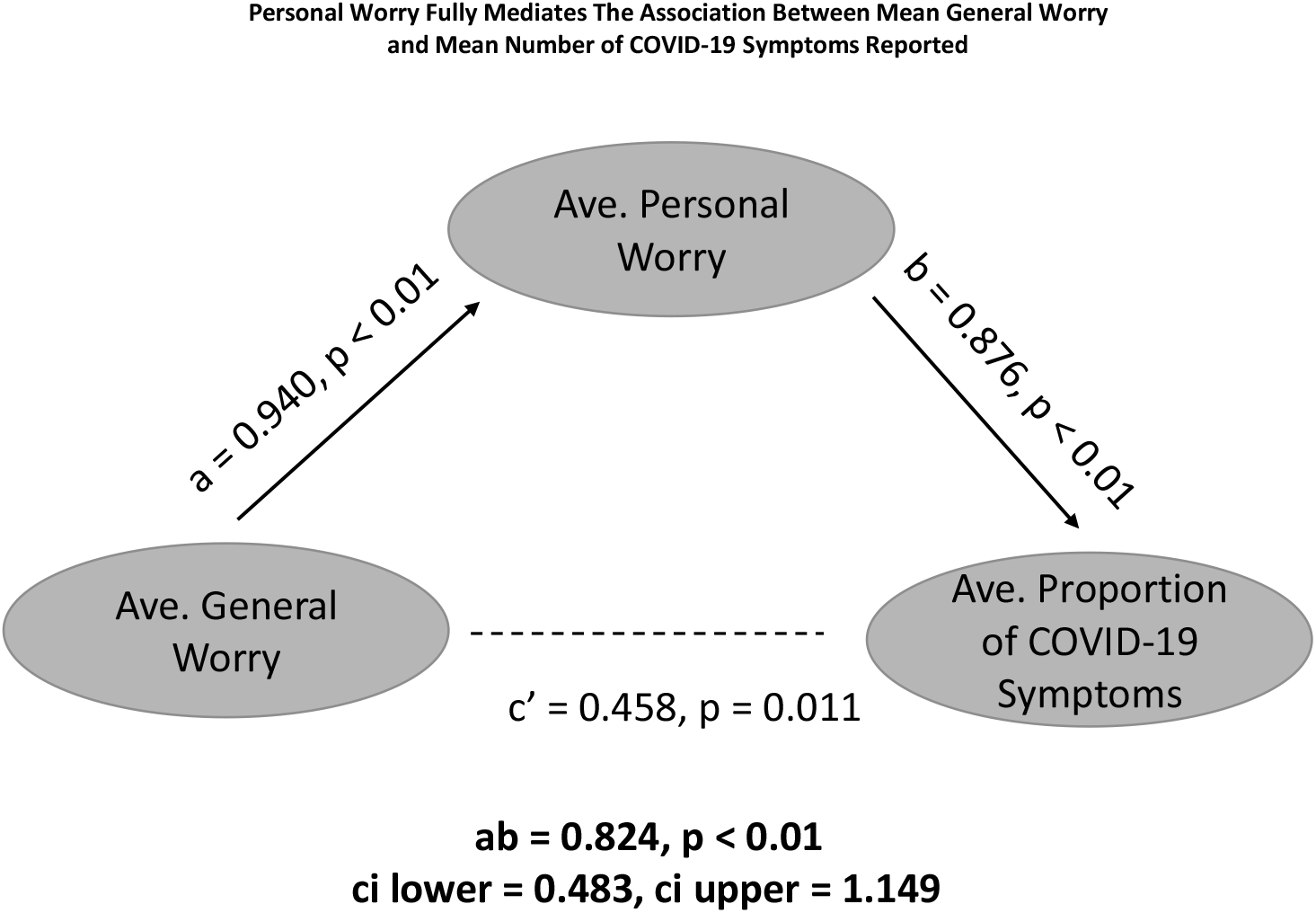
Mediation analysis shows that Personal Worry partially mediates the relationship between General Worry and symptoms are reported (indirect effect ab = 0.824, ci lower = 0.483, ci upper = 1.149). Direct effect of General worry on symptoms remains significant (c’ = 0.458, p = 0.011)

### Worried individuals have stronger positive associations between self-reported worries and subsequent symptoms

To build on our between-subjects analyses, we used within-subjects time-lagged analyses to test the hypothesis that worry at one time (T) would predict symptoms at the subsequent timepoint (T+1). Within subjects, we did not observe a significant main effect of participants’ general worry at any timepoint T (p>0.5), nor a moderation effect of mean general worry (p = 0.21) on the proportion of COVID-19 symptoms reported at T+1 (p = 0.63) (see Fig.3. a). We found that mean general worry (between subject effect) at time (T) was significantly associated with the average proportion of symptoms reported (i.e. the intercept; ß = 1.02, p<0.01), in line with our single-level regression analyses.

**Fig. 3 A:**
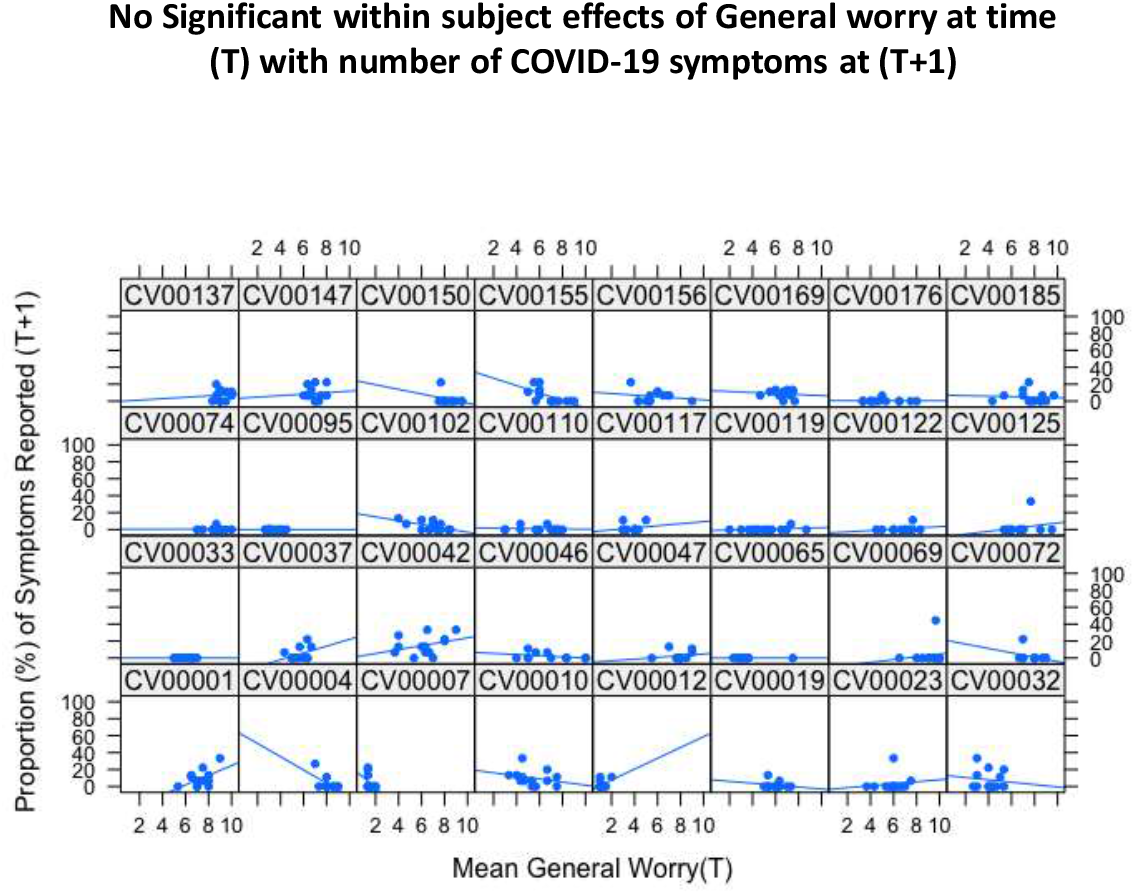
Example plots (n = 32) of Individual Participant relationships between Mean General Worry at each Timepoint (T) plotted against their reported Proportion of Symptoms at the next timepoint (T+1). Time –lagged analyses showed no significant within -subject associations between worry at T with subsequent symptom reporting at (T+1) (Table 5.i.)

**Fig. 3 B:**
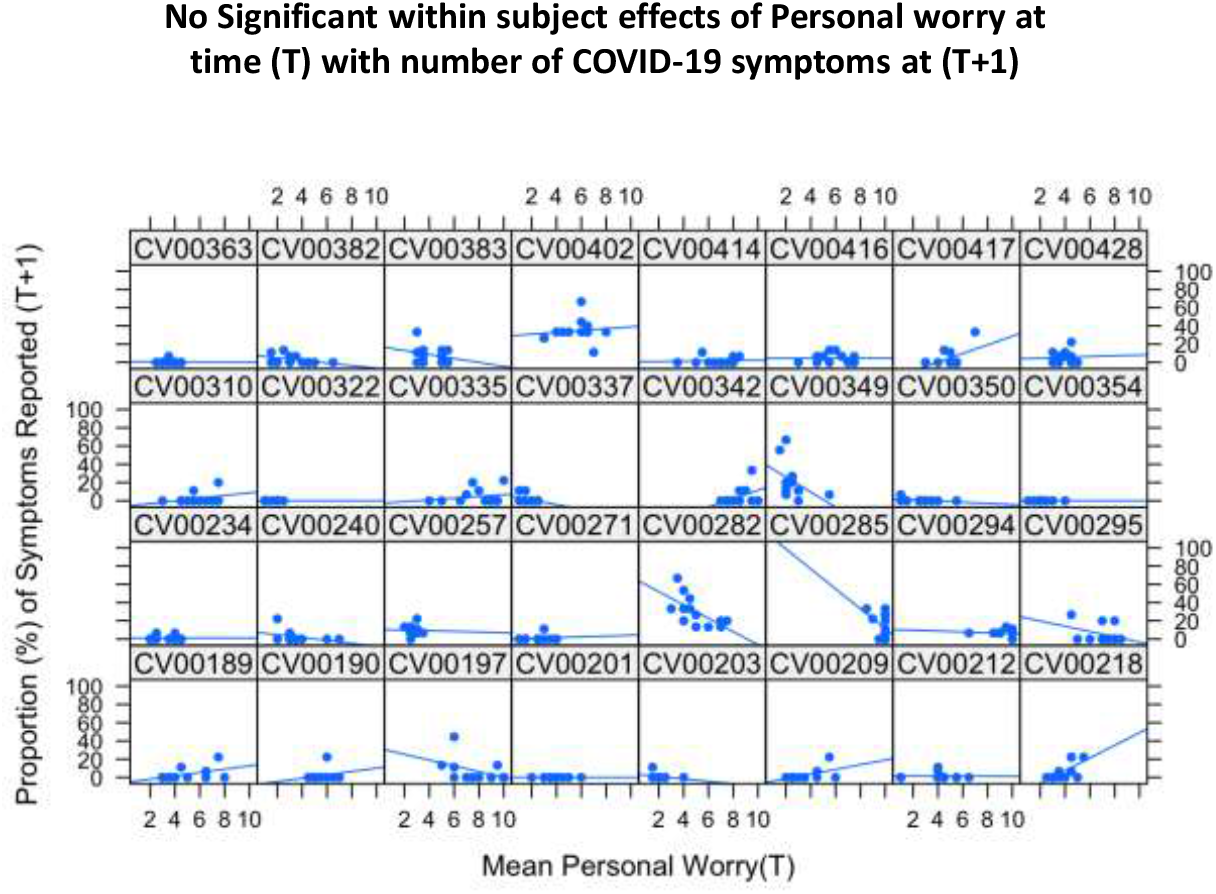
Example plots (n = 32) of Individual Participant relationships between Mean Personal Worry at each Timepoint (T) plotted against their reported Proportion of Symptoms at the next timepoint (T+1). Time –lagged analyses showed no significant within -subject associations between worry at T with subsequent symptom reporting at (T+1) (Table 5.ii)

Similar to general worry, we did not observe any association between personal worry and subsequent symptoms, whether we analyzed effects within subjects (p > 0.5) or evaluated potential moderation (p > 0.6). Consistent with the single level regressions, we did observe positive associations between mean personal worry and mean symptoms reported (ß = 0.91, p < 0.01).

## DISCUSSION

The COVID-19 pandemic represents a natural experiment as to how the population at large is affected by associated uncertainty, appraisal of risk and threats to health and daily life. Our results show that worries and negative expectations are associated with the experience of more COVID-19 related symptoms in people who do not report coronavirus infection during the first year of the pandemic. We found that increased general worries were associated with reporting more COVID-19 symptoms and that this association was mediated by worries about personal worries. We also found that mental health status, i.e., PPS, and the number of medical comorbidities were significant predictors for increased COVID-19 symptom reporting. However, our time-lagged analyses did not reveal associations between worries at one time and symptoms at the following timepoint, and thus we did not find evidence for postulated nocebo effects. Here we discuss these findings and their implications.

We assessed worry as a proxy for negative expectations that have been shown to modulate health outcomes (Hirsh 2015, Di Blasi 2001). Our delineation of worry measures into general and personal worry acknowledged a possible conceptual difference between the effects of outward expectations and self-directed expectations. As seen with hope and other cognitive constructs, changes in these different worry domains might have separate effects on personal well-being (Wiles 2008, Lee 2014). While the nocebo effect is thought to be associated with generalized negative expectations and anxiety, it is possible that different kinds of worry may be associated differentially with a nocebo effect. The partial mediation effect of personal worry reflects that other possible mediating factors exist, likely not captured in our models. However, it also highlights the relevance of health anxiety (as a sub-domain of generalized worry) in the formation of nocebo effects.

Importantly, anxiety about personal health has also been indicated as one of the relevant psychological factors causing people to *believe* they are infected with COVID-19 (Daniali 2021). These findings may suggest some added nuance in the relationship between negative expectations about health outcomes (which might include overall wellbeing, separate from contracting the viral infection) and nocebo effects.

The number of other health conditions (comorbidities) and the probability that participants had mental health conditions (PPS) were also significant predictors of increased COVID-19 symptom reporting. The former finding supports lines of research into one of the psychological mechanisms of nocebo effects—the misattribution of negative symptoms from pre-existing or unrelated comorbidities (Planes 2016). It is, however, also conceivable that somatic symptoms due to other medical conditions may be similar to or overlap with COVID-19 related symptoms. Furthermore, the finding on mental health status as a positive predictor is in line with previous research indicating that people with psychological symptoms such as anxiety and depression are more predisposed to developing nocebo effects (Davies 2003; Wells, Kaptchuk 2012; Planes 2016).

Although only a small percentage of our study sample was recruited from a pool of past NIMH study participants, our study was advertised by NIMH and mental health social networks, and thus our sample likely over-represents the prevalence of mental health conditions. If individuals with mental health conditions are predisposed to nocebo effects, further studies in populations without mental health conditions are necessary.

Our current results did not show any interaction effects between mental health status and the experience of general or personal worry on nocebo effects. However, the effect of mental health status and its possible interaction with worry and state vs trait anxiety is still important to consider in other cohorts given the associations of anxiety (Benedetti, 2006) and other mental health disorders effects (Planes 2016) with nocebo effects.

We also observed a small but significant association between age and reported symptoms, driven by younger participants reporting more symptoms. Though the effect is almost negligible, it is worth noting that this finding is in line with another study that also showed that younger age was related to more reports of COVID symptoms in a Norwegian anonymous sample (Daniali 2022). Research concerning age and nocebo effects is very sparse and we did not have any specific hypotheses regarding the effects of age and other demographic factors on symptom reporting. This finding adds to growing research showing that despite being at a comparatively lower risk for the most severe effects of COVID-19, younger individuals may experience greater effects of psychological distress during COVID-19 (American Psychological Association 2020, Horesh 2020). Though, we found significant interactions between personal worry and age and marginally significant interactions between personal worry and sex, however these effects were negligible (r<0.1) and thus are unlikely to be clinically meaningful in this sample. Some previous research does indicate that female sex is associated with increased reports of COVID-19 symptoms (Daniali 2022) and greater nocebo effects (Vambheim 2017). However, our sample cohort being predominantly female likely prevents us from appropriately addressing potential sex effects.

Arguably, the association between worry and symptoms experienced at any timepoint could be bidirectional (i.e. individuals with more symptoms may be more worried). To introduce the directionality needed to show a nocebo effect, we tested for any time-lagged effects of worries on symptom reporting. Within subjects, we found no significant time-lagged effects of general or personal worry at any timepoint on symptoms reported 2 weeks later and no interaction effects with individual average worry levels. In contrast, a recent article exploring similar nocebo effects found that a specific belief about COVID-19 symptom severity was positively associated with experiencing symptoms 3-4 weeks later (Rozenkrantz, 2022). However, participants from this study were studied at two timepoints after the removal of the first lockdown in Europe around May 2020. Since that was very early in the trajectory of the pandemic, other factors specific to that time window may have contributed to a nocebo effect. It is possible that at different critical time points doing the pandemic, people may be more prone to nocebo effects. However, our results, which span the entire first year of the pandemic in a larger sample (>500 participants versus 95-234 participants) with more frequent study intervals (12 responses every two weeks versus 1 response 3-4 weeks later) contribute to a more robust picture and a stronger test of whether nocebo effects had a large-scale impact on symptoms during the COVID-19 pandemic. In contrast to our hypotheses and findings from Rozenkrantz and colleagues, we found no causal evidence of the impact of worries on subsequent symptoms.

This study had several limitations that ought to be acknowledged. While we did exclude all participants who reported testing positive for COVID-19 at any timepoint during the survey period, the limited access to COVID-19 tests earlier on in the pandemic poses a limitation. It is possible that some participants may have reported symptoms that were actually due to coronavirus infection that was undetected and thus not a true nocebo effect. In addition, although we failed to find causal evidence of nocebo effects per se in time lagged analysis of biweekly reports of worries and symptoms, more fine-grained sampling (e.g. daily) may have detected that worries precede symptoms. Even if this were to be the case, a number of other factors would need to be assessed to isolate pure nocebo effects. Despite negative coronavirus tests, some participants may experience worry due to greater likelihood of exposure and infection, e.g., employment as an essential worker, family exposure, as well as elevated regional or community COVID-19 rates. Thus, studies that employ random assignment and directly manipulate beliefs and expectations about symptoms are necessary in order to truly measure the impact of nocebo on symptoms.

Because the COVID-19 pandemic is still ongoing and fast-changing, the psychological & health effects of the COVID-19 pandemic will likely be a subject to disentangle for a long time. While, our directional analyses do not show direct nocebo effects, these results recapitulate the association between worry and the experience of COVID-19 symptoms or potentially other adverse physical health outcomes, which may also be especially heightened during the context of a pandemic. With the current rise in health anxiety during the ongoing pandemic (Schimmenti 2020, Heinen 2021, Kibbey 2021) and given the recognized importance of more personalized healthcare, clinical care may benefit from assessments of worry and expectation factors. This has potential to help determine which patients might be more at risk for worse health outcomes during current or future pandemics. Interventions aimed at mitigating worries or managing expectations may also provide benefit to some patients.

Further studies investigating worry and symptoms in populations with confirmed negative COVID-19 tests or isolated populations will be needed to isolate the occurrence of true nocebo effects during the pandemic. As such, more empirical data to elucidate the psychological and behavioral factors associated with nocebo effects will be necessary to limit preventable negative health outcomes will help manage available resources. Further exploring the occurrence of nocebo effects and understanding populations that are susceptible to the effects of negative expectations, may inform healthcare management and help prevent worsening health outcomes.

## Data Availability

All data produced will be made available online through the NIMH Data Archives. Prior to that, data may be made available upon reasonable request to the authors.

## Acknowledgements

We would like to acknowledge and thank the following individuals for their statistical expertise and work in data management and cleaning of this huge study: Alison Gibbons, Cristan Farmer, PhD, Francisco Periera, PhD and Jacob Shaw.

## Statement of Ethics

All procedures were conducted in accordance with the Declaration of Helsinki and ethical approval was for this study was granted by the Institutional Review Board of the National Institutes of Health.

## Study approval statement

This study protocol was reviewed and approved by the Institutional Review Board of the National Institutes of Health.

## Consent to participate statement

All participants provided informed consent before their first entry into NIMH study participant pool.

## Author Contributions

All listed authors had complete access to the data in the study and were involved from the conception and design of the study.

