## Supplemental File for "The Effects of Expectations and Worries on the Experience of COVID-19 Symptoms"

**ABSTRACT**

***Introduction:*** The COVID-19 pandemic has been shown to have profound effects on both mental and physical health. Distress and widespread uncertainty about global events and personal risk are associated with increased worry and negative expectations that impact physical health. Thus, the current pandemic poses a possibility for the experience of nocebo effects.

***Objective:*** To evaluate the likelihood of nocebo-induced COVID-19 symptoms in a US sample.

***Methods:*** An online study on the mental health impact of COVID-19 asked participants to complete a set of biweekly surveys over a 6-month period between April 2020 and May 2021. We focus on responses from 3,027 individuals who reported never testing positive for COVID-19. We assessed the association between two types of worry and self-reported symptoms of COVID-19. We used multi-level models to examine variations across and within participants over time. We further investigated the effects of pre-existing health conditions and mental health status.

***Results:*** There was a positive association between symptoms and both general ($b= 2.56$, $p<0.01$) and personal worry ($b=2.77$, $p<0.01$). However, worry reported at one timepoint was not specifically associated with symptoms reported two weeks later ($p = 0.63$, $p=0.56$). We also found that a greater number of prior clinical comorbidities and greater mental health burden were significant predictors of symptom reporting.

***Conclusions:*** These results suggest that increased worries during the COVID-19 pandemic were associated with greater symptoms. Further studies investigating worry and symptoms in populations with confirmed negative COVID-19 tests or isolated populations will be needed to isolate the occurrence of true nocebo effects during the pandemic.

***Keywords***: Nocebo; COVID-19; Expectations; Mental health; Health Anxiety

| **Table 2.Pearson Correlations of Predictors and Outcome Variables** | | | | |
|---|---|---|---|---|
| | AGE | Female SEX | Number of Physical Comorbidities | Mental Health Burden Score (PPS) |
| **Average General Worry** | r = -0.042, p=0.02* | r = 0.14, p < 0.001*** | r = 0.104, p< 0.001*** | r = 0.258, p < 0.001*** |
| **Average Personal Worry** | r = 0.027, p=0.15 | r = 0.115, p< 0.001*** | r = 0.179, p< 0.001*** | r = 0.251, p< 0.001*** |
| **Average Proportion of COVID-19 Symptoms Reported** | r = -0.092, p<0.001*** | r = 0.1, p<0.001*** | r = 0.197, p<0.001*** | r = 0.333, p< 0.001*** |

**Baseline Study Measures**

***(Measures Completed Upon Study Enrollment*):**

-Demographics Questionnaires

-A Clinical History Checklist

- Functional Status via World Health Organization Disability Assessment Schedule II (WHODAS) Functional Status[12]

-Psychiatric and Family History via Family Interview for Genetic Studies Scale (modified FIGS)[7]

-The Alcohol Use Disorder Identification Test (AUDIT)[1]

- Drug Use (DSM- 5 Level 2 Substance Use – Adult)[8]

**Repeated / Biweekly Measures:**

***(Measures Completed at Enrollment and Biweekly throughout Study duration*):**

-Kessler-5 Distress Scale[5]

- Loneliness Measure via the (3-Item Loneliness Scale)[4]

-DSM-5 Self-Rated Level 1 Cross-Cutting Symptom Measure- Adult[6]

- Study-specific COVID-19-related responses and circumstances survey (Psychosocial Impact of COVID-19 survey (Full survey here: https://tools.niehs.nih.gov/dr2/index.cfm/resource/22587).

**End of Study Measures:**

***(Measures Completed at Final Timepoint)***

- Chronic Pain Graded Scale (CPGS)[13]

-Brief Trauma Questionnaire (BTQ)[10]

-Social Support (PROMIS Instrumental Support)[3]

-Emotional Support (PROMIS Emotional Support)[3]

-Personality Survey (BFI-10)[9]

-Motivation Survey (BISBAS)[2]

- Brief Resilience Scale (BRS)[11]
